# Deep Learning for Transesophageal Echocardiography View Classification

**DOI:** 10.1101/2023.06.11.23290759

**Authors:** Kirsten Steffner, Matthew Christensen, George Gill, Michael Bowdish, Justin Rhee, Abirami Kumaresan, Bryan He, James Zou, David Ouyang

**Affiliations:** Department of Anesthesiology, Perioperative and Pain Medicine, Stanford University; Department of Cardiology, Smidt Heart Institute, Cedars-Sinai Medical Center; Department of Cardiac Surgery, Smidt Heart Institute, Cedars-Sinai Medical Center; Department of Anesthesiology, Cedars-Sinai Medical Center; Department of Computer Science, Stanford University; Department of Biomedical Data Science, Stanford University

**Author notes:** Contact, 300 Pasteur Drive, Room H3580, Stanford, California 94305-5640.

## Abstract

Transesophageal echocardiography (TEE) imaging is a vital monitoring and diagnostic tool used during all major cardiac surgeries, guiding perioperative diagnoses, surgical decision-making, and hemodynamic evaluation in real-time. A key limitation to the automated evaluation of TEE data is the complexity and unstructured nature of the images, which demonstrate significant heterogeneity across varied views in the evaluation of different cardiac structures. In this study, we describe the first machine learning model for TEE view classification. We trained a convolutional neural network (CNN) to predict standardized TEE views using labeled intraoperative and intraprocedural TEE videos from Cedars-Sinai Medical Center (CSMC). We externally validated our model on intraoperative TEE videos from Stanford University Medical Center (SUMC). Accuracy of our model was high across all labeled views. The highest performance was achieved for the Trans-Gastric Left Ventricular Short Axis View (area under the receiver operating curve [AUC] = 0.971 at CSMC, 0.957 at SUMC), the Mid-Esophageal Long Axis View (AUC = 0.954 at CSMC, 0.905 at SUMC), the Mid-Esophageal Aortic Valve Short Axis View (AUC = 0.946 at CSMC, 0.898 at SUMC), and the Mid-Esophageal 4-Chamber View (AUC = 0.939 at CSMC, 0.902 at SUMC). Ultimately, we demonstrate that our unique deep learning model can accurately classify standardized TEE views, which will facilitate further downstream analyses for intraoperative TEE imaging.

## Introduction

Cardiovascular disease is a leading cause of death and disability worldwide and has been one of the top ten most important drivers of increasing global disease burden in the last three decades.^1^ Echocardiography is the most commonly used imaging modality in the assessment of cardiac structure, function, and disease.^2,3^ Despite the growth seen in the application of artificial intelligence (AI) to transthoracic echocardiography (TTE)^4–7^, the application of AI and machine learning to transesophageal echocardiography (TEE) remains relatively unexplored. Although more invasive, TEE imaging often offers higher resolution images and is particularly valuable as a monitoring and diagnostic tool in the management of cardiac surgery patients and patients undergoing transcatheter procedures for structural heart disease.^2,8,9^ As the standard of care, intraoperative TEE imaging is performed during all major cardiac surgeries, especially those requiring an open sternotomy and cardiopulmonary bypass (CPB), to help make perioperative diagnoses, guide surgical decision-making, and evaluate hemodynamic states in real-time.

Advances in AI for medical imaging demonstrate that machine learning models can be trained to classify human disease states^10^, identify phenotypic data^11–13^, and predict clinical outcomes with accuracy that outperforms clinical experts and more traditional clinical prediction models.^13–17^ The early, landmark literature in AI for medical imaging focused on two-dimensional, static images such as chest x-rays and pathology images. More recently, deep learning techniques have been applied to two- and three-dimensional images over time, such as echocardiography videos.^5,11,18,19^

Given the critically important role that TEE imaging plays in the evaluation of complex cardiovascular disease states and in the perioperative management of high-risk cardiac surgery patients, there is great potential value to be extracted from TEE images with advanced deep learning methodologies. The first step in the interpretation of any echocardiography video is to classify the view, to orient the observer to the anatomy and potential pathology contained within the image. Therefore, in this study we tested whether it was possible to train a convolutional neural network (CNN) to accurately classify eight standardized TEE views using labeled intraoperative and intraprocedural TEE images.

## Methods

### Cohort Selection and Data Processing

We obtained TEE image data for 2,144 randomly selected patients who underwent an intraoperative or intraprocedural TEE exam at Cedars-Sinai Medical Center (CSMC) between the years of 2016 and 2021. This resulted in 3,103 TEE videos, including intraoperative echocardiography images from open (via sternotomy) cardiothoracic surgical operations and intraprocedural echocardiography images from transcatheter procedures for structural heart disease. We also obtained TEE image data from randomly selected adult (age 18 years and older) patients who underwent an intraoperative TEE exam during open cardiothoracic surgery at Stanford University Medical Center (SUMC), resulting in an additional 465 TEE videos for an external test set. The Institutional Review Board at Cedars-Sinai Medical Center and the Institutional Review Board at Stanford University Medical Center both granted ethical approval for this study.

Guidelines established by the American Society of Echocardiography and the Society of Cardiovascular Anesthesiologists identify twenty-eight different TEE views necessary to complete a comprehensive intraoperative multi-plane TEE exam.^20^ In actual clinical practice, individual patient factors, anatomic variations and pathology, and time constraints can preclude the acquisition of all twenty-eight views. For our multi-category deep learning view classification model, we chose the eight most consistently acquired TEE views in the intraoperative assessment of cardiac surgery patients, including: the Mid-Esophageal (ME) 2-Chamber View, ME 4-Chamber View, ME Aortic Valve (AV) Short Axis (SAX) View, ME Bicaval View, ME Left Atrial Appendage View, ME Long Axis View, Trans-Gastric (TG) LV SAX View, and Aortic View. Four of our eight chosen views (ME 2-Chamber, ME 4-Chamber, ME AV SAX, ME Long Axis) represent pooled categories that include two standardized views that capture overlapping structures. We also chose to generalize two categories (the TG LV SAX and the Aortic Views), in order to increase the sample sizes of these classes. Ultrasound image data was converted from Digital Imaging and Communications in Medicine (DICOM) format data to AVI videos prior to machine learning analysis. All images and their associated metadata were de-identified prior to labeling, model training, and analysis.

### AI Model Design and Testing

We trained a CNN with residual connections and spatiotemporal convolutions using the R2+1D architecture^21^ to classify TEE views. Model weights were randomly initialized. Models were trained to minimize the cross entropy between the predicted view and the actual labeled view. We used an Adam optimizer^22^, a learning rate of 0.001, and a batch size of 44. We employed early stopping to cease model training after no further improvement on the validation set occurred. Our final model trained for nine epochs. The model was trained on 32-frame sub-clips of videos in the training set, with a temporal stride of two, yielding a final model input length of 16 frames. The starting frame of these sub-clips within their parent clips were randomized during training as a form of data augmentation. All model training was done using the Python library PyTorch. Our code is available online at https://github.com/echonet/tee-view-classifier.

All TEE videos were labeled by a single board-certified echocardiographer. An active learning approach was used to reduce the number of human annotations required. A classifier was initially trained on 500 randomly selected labeled TEE videos. With the partially trained model, inference was performed on unlabeled TEE videos, which were then categorized into buckets based on predicted view. In the next round of video labeling, the echocardiographer focused on the uncommon views and poorly performing classifications, and the model was subsequently retrained based on the additional labeled TEE videos. This iterative active learning approach was performed for five rounds until model performance was adequate. Only the training and validation sets were constructed with this active learning approach. The videos in both the internal and external test sets were independent and never seen during training.

### Statistical Analysis

An internal hold-out test dataset from CSMC which was never seen during model training was used to assess model performance. An external test set from SUMC was also used for additional validation and was never seen during model training. Model performance was assessed via AUROC. Two-sided 95% confidence intervals using 1,000 bootstrapped samples were computed for each calculation. Unsupervised t-Distributed Stochastic Neighbor Embedding (t-SNE) was used for clustering analysis^23^. Statistical analysis was performed in Python.

## Results

We trained a CNN to classify eight standardized TEE views. Our training and validation sets contained 2,600 unique videos (split 4:1), representing 2,036 patients. The model was tested on 503 randomly selected videos from CSMC and 465 randomly selected videos from SUMC, none of which were seen during model training. Characteristics of our training, validation, and test patients are shown in Table 1. Our datasets included a broad spectrum of anatomic variation, clinical pathology, and imaging indications representing the cardiac open surgical and transcatheter procedural populations seen at CSMC and SUMC (Table 2). The images also included a wide range of technical variation, including differences in spatial and temporal resolution, field of view depth and sector width, gain, image quality, and use of color flow Doppler (Figure 1).

**Table 1.** Clinical characteristics represented in the training, validation, and internal test data sets.

|  | Total | Train | Validation | Internal Test |
| --- | --- | --- | --- | --- |
| Number of videos | 3,103 | 2,076 | 524 | 503 |
| Age | 67.8 ($\pm 15.4$) | 67.9 ($\pm 15.4$) | 68.3 ($\pm 15.7$) | 66.9 ($\pm 15.2$) |
| White, n (%) | 2,223 (71.6%) | 1,496 (72.1%) | 365 (69.7%) | 362 (72.0%) |
| Black, n (%) | 317 (10.2%) | 224 (10.8%) | 62 (11.8%) | 31 (6.2%) |
| Other/Unknown, n (%) | 271 (8.7%) | 177 (8.5%) | 41 (7.8%) | 53 (10.5%) |
| Asian, n (%) | 242 (7.8%) | 152 (7.3%) | 40 (7.6%) | 50 (9.9%) |
| Pacific Islander, n (%) | 35 (1.1%) | 18 (0.9%) | 14 (2.7%) | 3 (0.6%) |
| Native American, n (%) | 12 (0.4%) | 6 (0.3%) | 2 (0.4%) | 4 (0.8%) |
| Hispanic ethnicity, n (%) | 395 (12.7%) | 255 (12.3%) | 68 (13.0%) | 72 (14.3%) |
| Female gender, n (%) | 1,120 (36.1%) | 757 (36.5%) | 194 (37.0%) | 169 (33.6%) |
| Atrial fibrillation, n (%) | 1,146 (36.9%) | 773 (37.2%) | 194 (37.0%) | 179 (35.6%) |
| Heart failure, n (%) | 1,504 (48.5%) | 1,009 (48.6%) | 263 (50.2%) | 232 (46.1%) |
| Hypertension, n (%) | 1,681 (54.2%) | 1,142 (55.0%) | 267 (51.0%) | 272 (54.1%) |
| Diabetes mellitus, n (%) | 702 (22.6%) | 483 (23.3%) | 112 (21.4%) | 107 (21.3%) |
| Ischemic stroke, n (%) | 365 (11.8%) | 255 (12.3%) | 53 (10.1%) | 57 (11.3%) |
| Transient ischemic attack, n (%) | 194 (6.3%) | 137 (6.6%) | 31 (5.9%) | 26 (5.2%) |
| Pulmonary embolism, n (%) | 90 (2.9%) | 63 (3.0%) | 21 (4.0%) | 6 (1.2%) |
| Myocardial infarction, n (%) | 339 (10.9%) | 214 (10.3%) | 55 (10.5%) | 70 (13.9%) |
| Peripheral artery disease, n (%) | 540 (17.4%) | 376 (18.1%) | 83 (15.8%) | 81 (16.1%) |
| Vascular disease, n (%) | 814 (26.2%) | 551 (26.5%) | 126 (24.0%) | 137 (27.2%) |
| Coronary artery disease, n (%) | 1,152 (37.1%) | 769 (37.0%) | 192 (36.6%) | 191 (38.0%) |
| Chronic kidney disease, n (%) | 745 (24.0%) | 495 (23.8%) | 140 (26.7%) | 110 (21.9%) |
| Liver disease, n (%) | 159 (5.1%) | 110 (5.3%) | 24 (4.6%) | 25 (5.0%) |
| Chronic obstructive pulmonary disease, n (%) | 200 (6.4%) | 139 (6.7%) | 27 (5.2%) | 34 (6.8%) |
| Prior smoker, n (%) | 181 (5.8%) | 119 (5.7%) | 35 (6.7%) | 27 (5.4%) |

**Table 2.** Surgery or procedure types represented in our training, validation, and internal test data sets. CABG = Coronary Artery Bypass Graft.

|  | Total | Train | Validation | Internal Test |
| --- | --- | --- | --- | --- |
| Number of videos | 3,103 | 2,076 | 524 | 503 |
| CABG (%) | 194 (6.3) | 124 (6.0) | 31 (5.9) | 39 (7.8) |
| Valve procedure (%) | 321 (10.3) | 215 (10.4) | 42 (8.0) | 64 (12.7) |
| Aortic procedure (%) | 42 (1.4) | 30 (1.4) | 3 (0.6) | 9 (1.8) |
| Combination of CABG, valve, and/or aortic procedure | 218 (7.0) | 140 (6.7) | 35 (6.7) | 43 (8.5) |
| Other open cardiac procedure (%) | 317 (10.2) | 215 (10.4) | 56 (10.7) | 46 (9.1) |
| Mechanical circulatory support (%) | 52 (1.7) | 30 (1.4) | 13 (2.5) | 9 (1.8) |
| Transcatheter procedure (%) | 1959 (63.1) | 1322 (63.7) | 344 (65.6) | 293 (58.3) |

**Figure 1.**
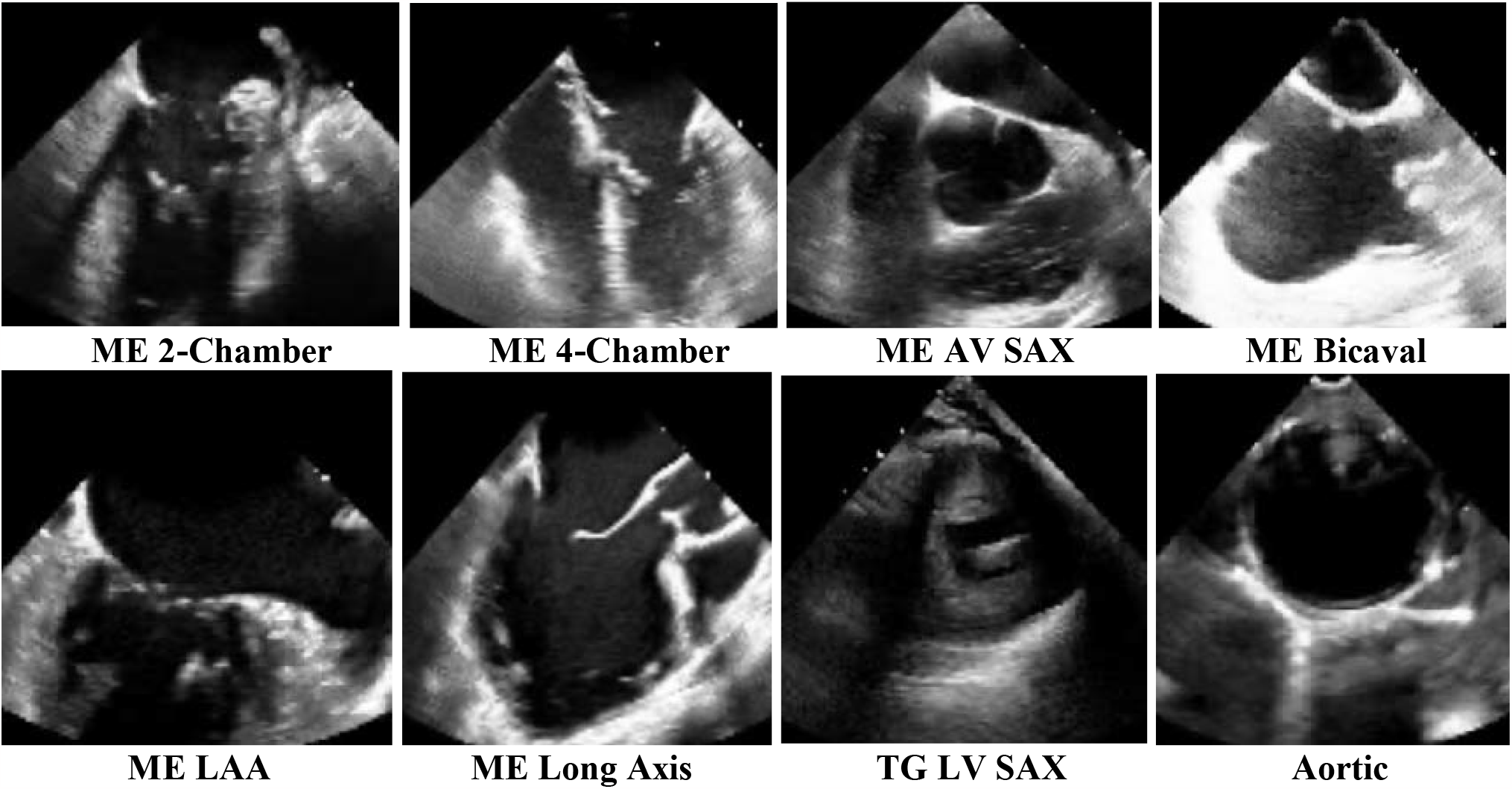
Sample training images used for the deep-learning view classification task. Above images are 2-dimensional still frames sampled from the video data used in model training. Eight standard TEE views were chosen, including: the ME 2-Chamber View, ME 4-Chamber View, ME AV SAX View, ME Bicaval View, ME LAA View, ME Long Axis View, TG LV SAX View, and Aortic View. TEE = transesophageal echocardiography; ME = mid-esophageal; AV = aortic valve; SAX = short axis; LAA = left atrial appendage; TG = trans-gastric; LV = left ventricular.

Our view classification model achieved an overall micro-averaged area under the receiver operating curve (AUC) of 0.919 on a hold-out CSMC test set of TEE videos (Figure 2). Our model showed particularly good performance for the Trans-Gastric Left Ventricular Short Axis View (AUC = 0.971), the Mid-Esophageal Long Axis View (AUC = 0.954), the Mid-Esophageal Aortic Valve Short Axis View (AUC = 0.946), and the Mid-Esophageal 4-Chamber View (AUC = 0.939).

**Figure 2.**
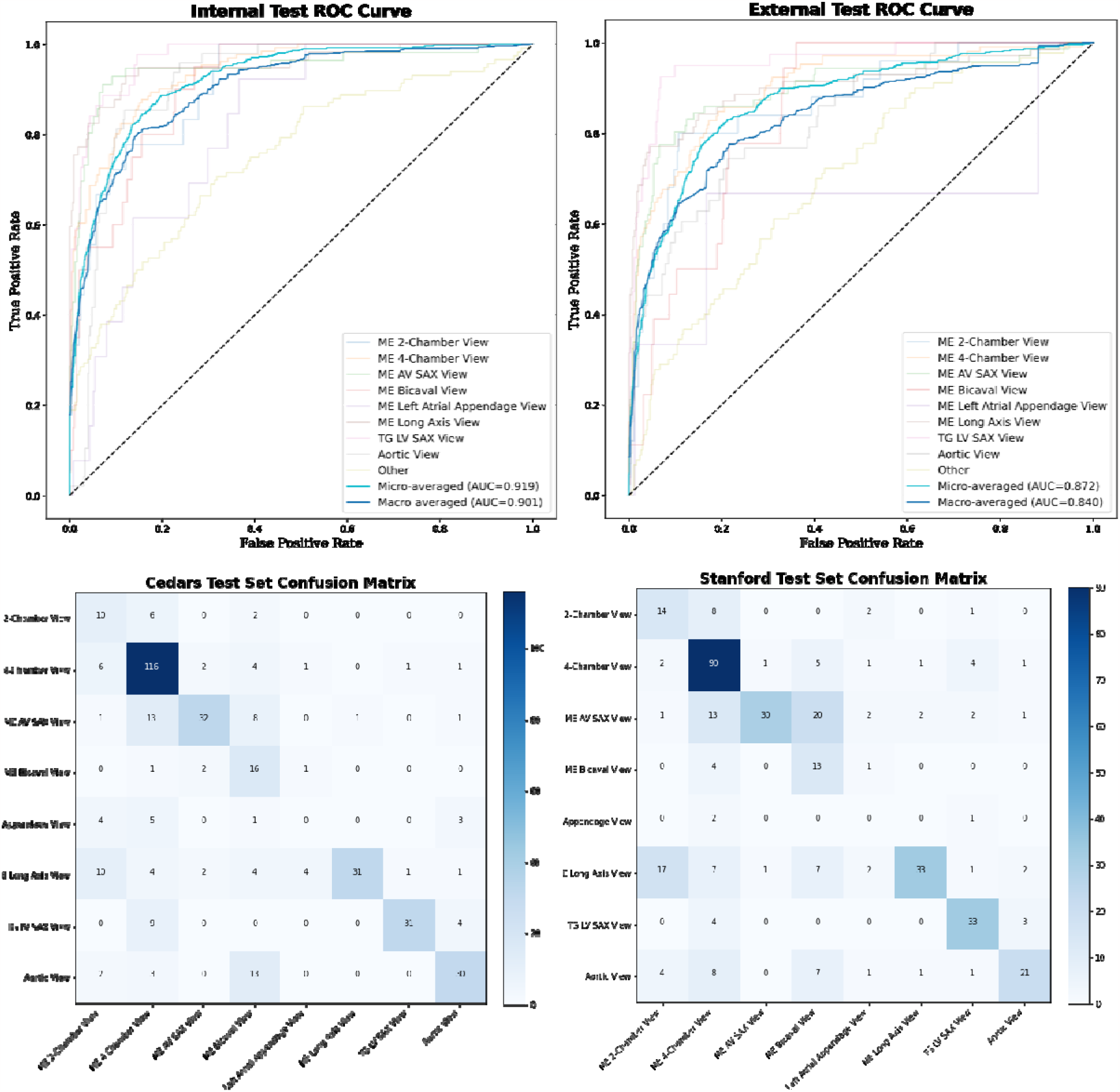
View classification model performance on internal (CSMC) hold-out test set and external (SUMC) test set. (a) AUC’s for each view class, demonstrating high accuracy (with AUC’s ranging from 0.816 – 0.957). No AUC was able to be calculated for the ME Left Atrial Appendage View in the randomly selected SUMC test set due to low sampling. (b) Confusion matrices showing model performance, with views labeled by a board-certified echocardiographer along the vertical axis and views predicted by the deep learning model on the horizontal axis. Numerical values in the matrices represent the number of images with the indicated ground-truth and model-predicted labels. Color intensity on the heatmap represents model accuracy. AUC = area under the receiver operating curve; CSMC = Cedars Sinai Medical Center; SUMC = Stanford University Medical Center; ME = mid-esophageal; AV = aortic valve; SAX = short axis; TG = trans-gastric; LV = left ventricular.

The model performance also generalized well externally, achieving a micro-averaged AUC of 0.872 when tested on 465 never-before-seen TEE videos from SUMC. Our model had similar performance for the Trans-Gastric Left Ventricular Short Axis View (AUC = 0.957), the Mid-Esophageal Long Axis View (AUC = 0.905), the Mid-Esophageal Aortic Valve Short Axis View (AUC = 0.898), and the Mid-Esophageal 4-Chamber View (AUC = 0.902) in the SUMC dataset.

Clustering analysis suggests our AI model can identify a meaningful embedding space representing the various TEE views from heterogeneous video input that generalizes across two institutions (Figure 3). Model performance was similar in standard black-and-white 2D B-Mode TEE videos (micro-averaged AUC = 0.902) and videos incorporating color flow Doppler information (micro-averaged AUC = 0.877) (Figure 4), the analyses for which were performed on a combination of the internal and external test videos due to the overall low prevalence of color flow Doppler videos in our datasets.

**Figure 3.**
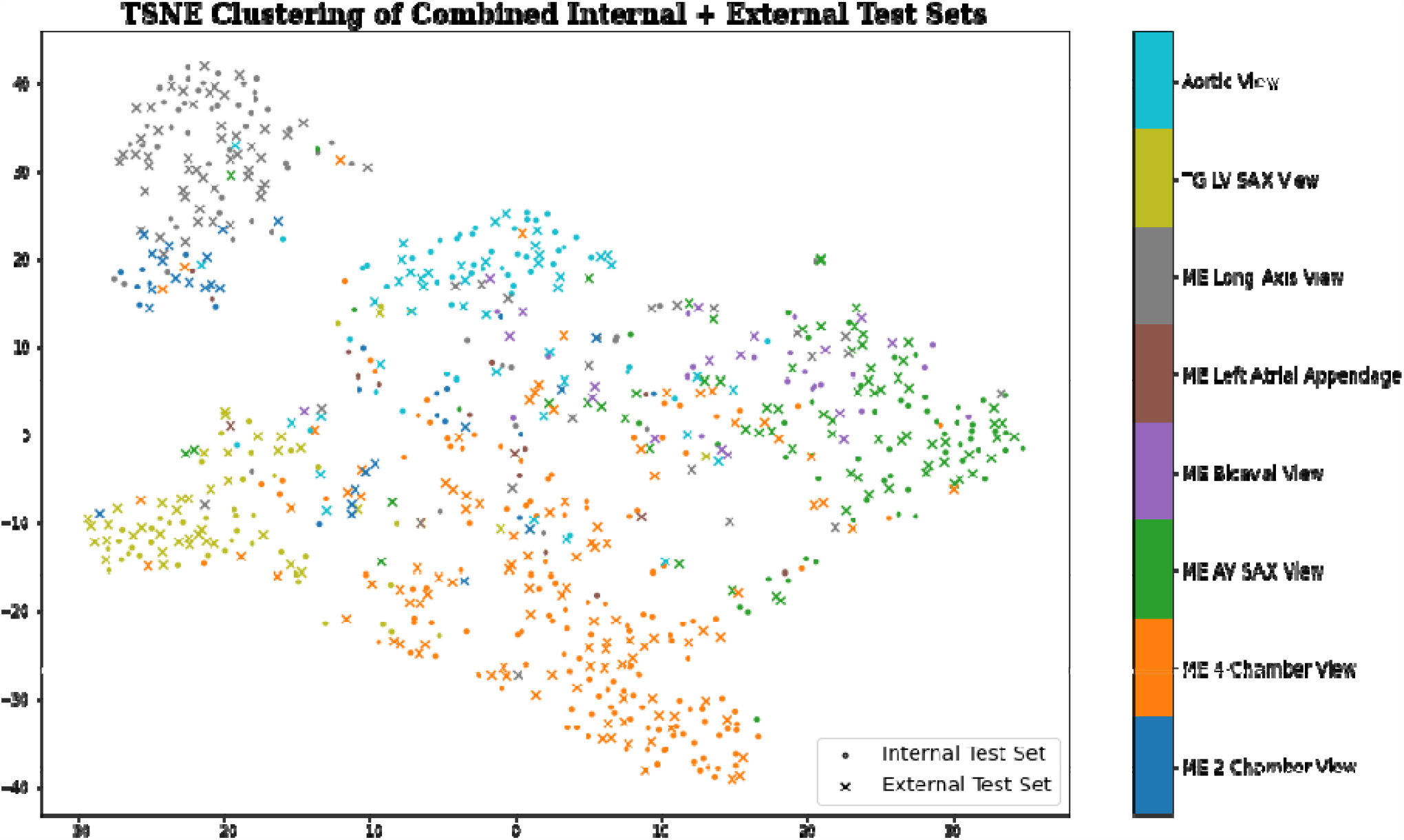
Clustering analysis showing the model’s ability to distinguish among standard TEE views. t-SNE clustering analysis of input images demonstrates that meaningful representations of standard TEE views are clustered appropriately together. In other words, images are sorted into groups that reflect standard TEE classes. Embedding representation is consistent across CSMC and SUMC, suggesting robustness and generalizability of the approach. TEE = transesophageal echocardiography; t-SNE = t-Distributed Stochastic Neighbor Embedding; CSMC = Cedars Sinai Medical Center; SUMC = Stanford University Medical Center.

**Figure 4.**
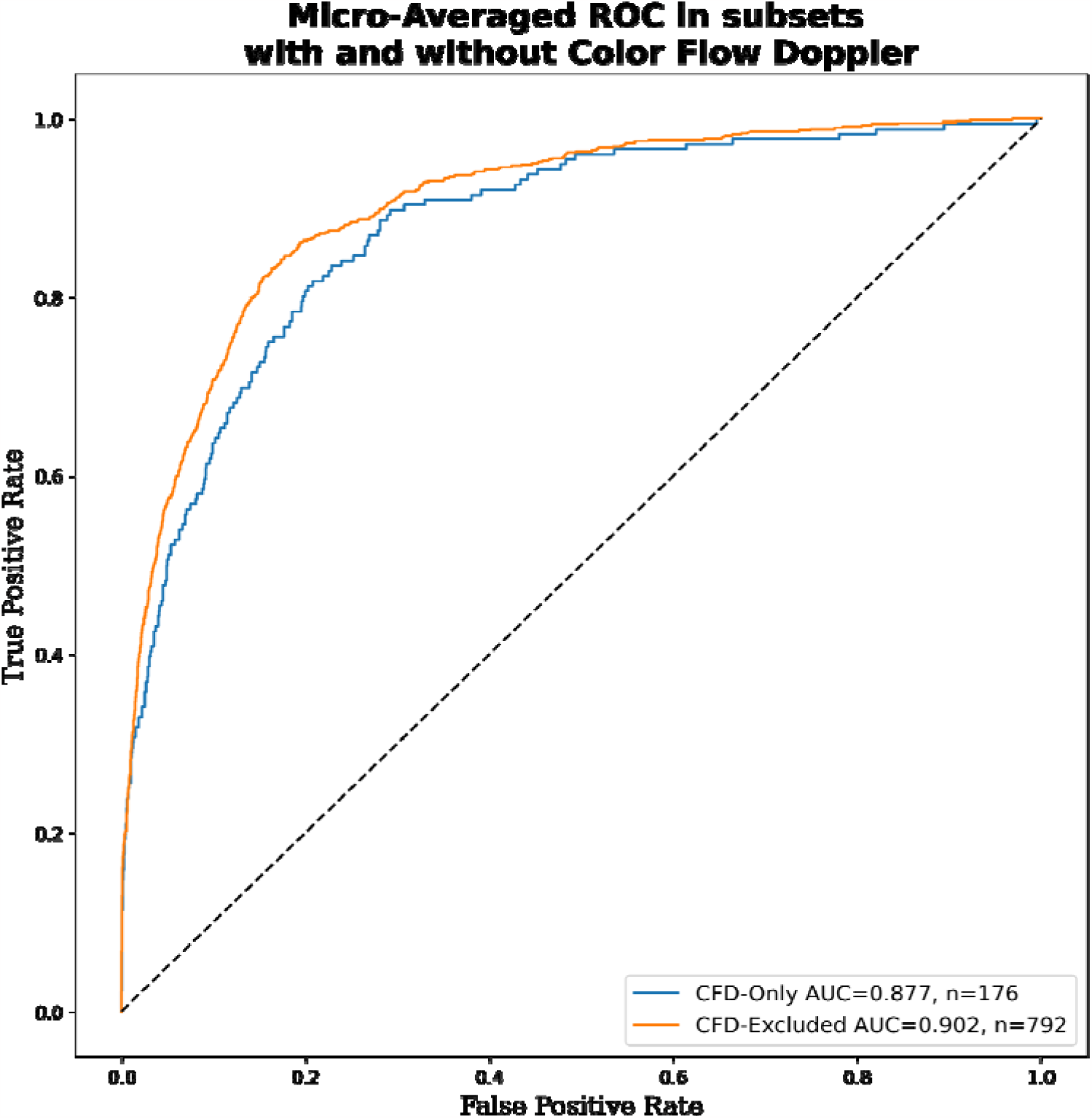
Micro-averaged receiver operating characteristic curves for model predictions in subset containing all color flow Doppler videos versus no color flow Doppler videos. This evaluation wa performed using a combination of the internal and external test sets due to the low prevalence of color flow Doppler videos in our data sets.

## Conclusions

Our deep learning model was able to classify the most commonly used intraoperative and intraprocedural TEE views with high accuracy across a wide range of clinical and echocardiographic characteristics. Our videos included patients undergoing many different types of open cardiac surgery and transcatheter procedures, representing a broad spectrum of anatomic pathology and differences in practice patterns across two institutions. The presence or absence of medical devices in our images (prosthetic valves, procedural wires, pacemakers, MitraClips) was highly variable among our datasets. Images also varied with respect to resolution, sizing and focus of the field of view, and the use of color flow Doppler. The model performance was consistent across the range of findings in both held-out internal and external test datasets, demonstrating the generalizability of our view classifier in real-world clinical contexts.

Our study represents the first application of a machine learning strategy to TEE videos. Prior AI-driven echocardiography studies focused primarily on TTE videos. It has been demonstrated that machine learning algorithms can be trained to identify standard TTE views from labeled datasets.^18,19,24^ Subsequent studies were able to take advantage of the standard clinical workflow for transthoracic imaging, which incorporates anatomic tracings and quantitative measurements, in order to streamline segmentation and classification tasks.^4,7^ It has also been shown that machine learning algorithms trained on TTE videos are able to recognize cardiac structures, approximate cardiac function, make accurate diagnoses, identify phenotypic information that is otherwise not easily recognized by a human observer, and predict clinical outcomes.^6,7,10,11,25^ Without an automated preprocessing and view classification pipeline for TEE, the ability to perform downstream deep learning tasks for TEE remained challenging.

The greatest barrier to applying AI to intraoperative and intraprocedural TEE imaging is the relatively unstructured nature of TEE data. TEE data is inherently variable because the environment in the cardiac surgery operating rooms is highly dynamic, which results in the acquisition of varying image sequences, non-standard views, and missing views. Moreover, intraoperative TEE exams are subject to significant clinical variation within a single study and across studies (changes in cardiac loading conditions, on-versus off-CPB, changes secondary to surgical manipulation, pre-versus post-surgical intervention, pharmacologic interventions, external cardiac pacing). However, given the significant role that TEE plays in the management of complex cardiac pathology and high-risk surgical patients, attempting to extract additional value from TEE images via deep learning strategies is worthy of exploration. The present study represents the first deep learning-based TEE view classification model trained on TEE videos.

The development of a view classification model for TEE images will extend the application of deep learning strategies to intraoperative echocardiography imaging. Previous work has already shown that intraoperative TEE imaging actively informs surgical decision-making^26,27^ and is associated with improved clinical outcomes after cardiac surgery.^28,29^ The development of AI-driven models based on intraoperative TEE images has the potential to further enhance the value of echocardiography in the perioperative and periprocedural period by improving the ability to diagnose cardiac surgical diseases and complications, diagnose the underlying etiology of varied hemodynamic states, and predict clinical outcomes in the immediate and long-term postoperative periods.

In summary, we show that an intraoperative and intraprocedural TEE-based deep learning model can accurately identify standardized TEE views, the first step in the AI interpretation of TEE images. Our study represents an important first step towards the automated evaluation of intraoperative imaging and the leveraging of deep learning strategies for the advancement of patient care.

## Data Availability

Our code is available online at https://github.com/echonet/tee-view-classifier.

